# Vitamin D status and cardiometabolic disease risk among healthy adults of Northern Ghana

**DOI:** 10.1101/2022.11.24.22282710

**Authors:** Abdul-Malik Bawah, Reginald A. Annan, Basma Ellahi, Karani SanthanakrishnanVimaleswaran, Abdul Rahman Haadi

**Affiliations:** Department of Biochemistry and Biotechnology, College of Science, Kwame Nkrumah University of Science and Technology, Kumasi, Ghana; Department of Nutrition and Dietetics, Faculty of Allied Health and Pharmaceutical Sciences, Tamale Technical University, Tamale, Ghana; Hugh Sinclair Unit of Human Nutrition, Department of Food and Nutritional Sciences, University of Reading, Reading RG6 6AP, UK; Hugh Sinclair Unit of Human Nutrition and Institute for Cardiovascular and Metabolic Research, University of Reading, Reading RG6 6AP, UK; Faculty of Health and Social Care, University of Chester, Riverside Campus, Chester CH1 4BJ, UK; Department of Statistical Sciences, Faculty of Applied Sciences and Technology, Tamale Technical University, Tamale, Ghana

**Author notes:** Correspondence (A.-M.B.). (A.-M.B.) (R.A.A.). (S.A.).

**Keywords:** Vitamin D, Risk factors, Cardiometabolic Disease, Healthy Adults

## Abstract

Cardiometabolic Disease (CMD) is a cluster of conditions that increase the risk for cardiovascular events, but its relationship with vitamin D status has not been studied in Ghana. A cross-sectional study design was conducted to investigate the relationship between serum vitamin D status and CMD risk (obesity, type 2 diabetes mellitus, hypertension, and dyslipidemia) among 302 apparently healthy adults, aged 25 to 59 and of northern decent in Ghana. Biochemical analysis was done to determine serum total cholesterol (TC), Triglycerides (TG), High Density Lipoprotein (HDL), Low Density Lipoprotein (LDL), Fasting Blood Glucose (FBG), Glycated Haemoglobin (HbA1c) and serum vitamin D levels of participants. Anthropometric assessment was also done and BMI, WC, HC, Blood Pressure, WHR, Body Fat (BF) and visceral fat percentages were obtained. Analysis was done using SPSS (version 25). We evaluated CMD risk factors to predict vitamin D status using binary and multiple linear logistics regression analysis. Similar between gender, participants’ mean age was 38.78years (SD 10.42years). The females had higher BMI (24.31 SD 7.96 versus 22.52 SD 3.07 kgm^2^, p=0.006), % total body fat (24.31 SD 7.96 versus 22.52 SD 3.07%, p=0.001), WC (24.31 SD 7.96 versus 22.52 SD 3.07 inches, p=0.002), and HC (24.31 SD 7.96 versus 22.52 SD 3.07, p=0.002 inches) than the males, while the males had higher mean total cholesterol (5.74 **SD** 1.02 versus 3.57 **SD** 1.02) and LDL (5.40 **SD** 1.05, verse 3.22 **SD** 1.09, p=0.005) than the females. Serum vitamin D levels was significantly associated with age (**p= 0.047**) but not gender (**p=0.349**). Overall, 6.4% of participants had CMD, representing 10.2% of the females, and no male. Multivariate analysis reveals gender, average BP, FBG, and % total body fat to significantly affect serum vitamin D concentrations. Seven percent (7%) of participants were serum vitamin D deficient (VDD) and 28.5% had serum vitamin D insufficiency (VDI), similar by gender, but increased with age (p=0.047). Both mean SBP (P<0.0001) and DBP (p<0.0001) increased with VDD, while means of FBS (p=0.032), BMI (p=0.004), HbA1c (p=0.040), and LDL (p=0.047) are highest in the VDD group and least in the Vitamin D Sufficient (VDS) group. Binary Logistic regression showed participants with high SBP (**OR= 0.055; 95% CI= 0.008-0.361; p= 0.003**) and high FBS (**OR= 0.076; 95% CI= 0.014-0.404; p= 0.002**) had increased odds of VDD compared with normal levels of SBP and FBS. Multiple logistics regression showed FBG, TG, LDL and WC significantly predicted CMD. In conclusion, CMD risk factors were common among the participants and more than a third were vitamin D deficient or insufficient. Individual CMD risk factors increased with vitamin D deficiency, but CMD did not predict serum vitamin D status.

## Introduction

Vitamin D is a fat-soluble vitamin that is mainly produced by the body when the human skin is exposed to the ultraviolet rays of the sun (Holick, 2008b). It is also obtained naturally as a nutrient from foods like mushroom that is sun-dried and oily fish. When vitamin D is obtained from diet or synthesis from sunlight, it enters the circulatory system and becomes hydroxylated to form 25-hydroxyvitamin D (25(OH) D) in the liver. Even though this circulatory form of vitamin D is not bio-active, its serum concentration is a reflection of the nutritional status of Vitamin D. Further hydroxylation of the 25(OH) D occurs in the kidneys to form the active metabolite known as dihydroxy vitamin D (1, 25(OH)2 D). The 1, 25(OH)2 D) molecule then binds to vitamin D receptors (VDR) in target cells forming a vitamin-receptor complex which expresses the biological effect of the vitamin.

Biologically, vitamin D acts on the kidneys, small intestines and bone to regulate calcium and phosphorus metabolism (DeLuca & Zierold, 1998). The 1, 25 (OH)2 D is synthesized in many tissues and the VDR is expressed throughout the body. This makes vitamin D very essential in many non-skeletal body functions, such as cardiovascular and immune functions (Holick, 2008a; Agmon-Levin et al., 2013; Luong & Nguyen, 2013). For this reason, vitamin D deficiency may be responsible for the pathological conditions of many Non-Communicable Diseases, including CVDs (Abu el Maaty & Gad, 2013), autoimmune diseases (Agmon-Levin et al., 2013), and some cancers (Sun et al., Hollis et al., 2013).

There is proof from animal and human studies that 1,25(OH)2D improves insulin sensitivity, restricts rennin synthesis and strengthens contractility of cardiac muscle, thereby promoting cardiac function (Belenchia et al., 2013; Kayaniyil et al., 2010; Matias et al., 2010; Vaildya & Williams, 2012; Xiang et al., 2005). Therefore, optimum vitamin D status can contribute to reducing the risk of cardiometabolic disease, including obesity, insulin resistance, hyperglycemia, dyslipidemia, and hypertension.

Cardiometabolic Disease (CMD) is a combination of risk factors that cause cardiovascular events. These include central obesity, hypertension, insulin resistance, hyperglycemia, decreased levels of serum high density lipoprotein-cholesterol (HDL-C), increased levels of serum Low Density Lipoprotein-cholesterol (LDL-C), and increased serum triglycerides (TGs) levels (Alberti et al., 2005). Different criteria exist for diagnosing CMD. However, central obesity and insulin resistance/impaired glucose tolerance or diabetes are central components of CMD. Although long-term unhealthy lifestyle is an important identified risk factor for CMD (Kwasniewska et al., 2009), some observational studies have associated hypovitaminosis D with CMD (Brock et al., 2010; González-Molero et al., 2013, Wamberg et al., 2013). Yet, the associations between vitamin D status and CMD from several Randomized Controlled Trials (RCT) have been inconsistent (Wood *et*., *al*, 2014; Petchey *et al*., 13; Salehpour *et al*., 2012; Von Hurst *et al*., 2010), and understudied in adult populations of Ghana.

Obesity, hypertension and dyslipidemia are on the rise globally and in Ghana. Studies in 2020 have reported overweight and obesity prevalence of more than 50% among Ghanaian adults (Agyapong et al., 2020) and increased CMD risk factors such as hypertension, dyslipidemia and physical inactivity (Agyapong et al., 2017). Essien et al., (2019) found overweight and hypertension among police officers in Ghana to be 40% and 34.2% respectively, with 65.8% having dyslipidemia (Miriam et al., 2019). Also, an association have been reported between obesity and genetic predisposition among Ghanaian adults in Kumasi (Alsulami et al., 2020), where higher intakes of total fat, SFA, MUFA and PUFA among genetically predisposed adults was associated with increased waist circumference. In Ghana, a study reported a positive correlation between vitamin D deficiency and Type 2 diabetes (T2DM) among Ghanaian type 2 diabetics seeking healthcare at the Nkawwie Government Hospital (Fondjo et al., 2017). Our study sought to investigate the association between vitamin D status and CMD among healthy Ghanaian adults. Understanding these relationships among Ghanaians is a step closer to addressing the rising CMD in many Low and Middle Income Countries (LMIC).

## Materials and Methods

### Study Design

A cross-sectional study design was used to investigate the relationship between serum vitamin D status and CMD risk of apparently healthy adults of Northern decent in Ghana.

### Study area

The study was conducted in the capital of the Northern Region of Ghana, Tamale among the Dagomba ethnic group. Ghana is located in West Africa, south of the Sahara and on the West of the Gulf of Guinea. The country is divided into 16 administrative regions with the Greater Accra Region as the national and administrative capital. According to the 2021 Population and Housing Census (PHC), the total population of Ghana was **30**,**832**,**019** with an annual growth rate of 2.5%. Tamale is the capital town of the Northern Region. It is located within the Guinea Savannah belt and is the fourth largest city in Ghana, with a population of 2, 310, 939 inhabitants who are predominantly Dagombas, and with a growth rate of 7.5% (GSS, 2021 PHC). The size of Tamale is approximately 922km sq. and is considered the fastest growing city in West Africa (http://www.ghanadistricts.com), Tamale is located 600 km north of <u>Accra</u>-the capital city of Ghana. The metropolis experiences one rainy season from April to September or October with a peak in July and August. The mean annual rainfall is 1100 mm within 95 days of rainfall in the form of tropical showers. Consequently, staple crop farming is highly restricted by the short rainy season. (GLSS, 2005/2006). The northern region is one of the poorest of the then three northern regions (now five) of Ghana (GLSS, 2005/2006).The population of adults in Northern Region aged 25 to 59 years according to the latest 2010 population was 741, 318 (GSS, 2010 PHC).

### Study Population

The study participants were selected from among free living and apparently healthy individuals aged 25-59 who resided in four selected communities of Tamale. The inclusion criteria were: (1) age> 25 years and < 60 years of age, (2) Ghanaian by ancestral origin and (3) Apparently healthy, defined as no physical complaints at the time of recruitment), no known communicable or non-communicable chronic diseases and cardiovascular disease. Pregnant women or people on lipid lowering drugs/ anti-diabetic drugs/ anti-hypertensive drugs were excluded.

### Recruitment and Sampling Technique

Three hundred adult volunteers, aged 25 to 59 years old, who were of northern extraction and Dagomba ancestry, and living in Tamale in the Northern Region were recruited according to the inclusion criteria. The participants were recruited from households in three communities in Tamale, namely Dungu, Dohani and Zujung communities. These three communities were randomly selected from a list of indigenous communities in Tamale. The individual participants were then selected by directly contacting individual adults and tracing them to their homes or going into households to identify adults who were willing to participate in the study. Each participant was informed of the study, and those who were willing to participate were screened for inclusion or otherwise using the screening interview questions. Screening was done by administering the screening interview question to participants. The questions included a set of inclusion and exclusion criteria stated above. Those who met the inclusion criteria were deemed to have qualified for participation. Any participant who passed the screening interview, was asked to observe 12 hours overnight fast for blood samples to be taken. The blood sample of the participants was taken early the next morning and participants completed the main questionnaires.

## Data collection

### Socio-demographic and lifestyle data

Data on socio-demographic characteristics and lifestyle were collected using a structured questionnaire. Data collected included alcohol intake, smoking, daily physical exercise and past medical history. For female respondents, contraceptive use and menopause were also elicited. History of chronic diseases and treatments of these chronic diseases, such as hypertension, diabetes mellitus and dyslipidemia were also elicited by the questionnaire.

### Assessment of Body Composition

For the purpose of obtaining accurate anthropometric measures, participants were encouraged to empty their bowels and bladder if they had the urge to do so before the anthropometric measures were taken. Also, only light clothing without shoes and cap/hats were accepted as prescribed dressing for the measurement of participants’ weight and height. Participants stood on the stadiometer in an upright posture while their feet were apposed and the height measures were read. The weights of participants were measured using the Body Composition Analyzer (BCA), which automatically calculated the Body Mass Index (BMI) after the height and age parameters of the participants were inputted into the BCA. To measure waist circumference, a tape measure was placed at the midpoint between the lowest rib and the iliac crest, as described by Klein et al., 2007. Three measurements were taken, and means were calculated to the nearest 0.1cm. The measurement of Hip Circumference (HC) was done using the widest point between the iliac crest and buttock as described by Bengtsson et al., (1993). Participants were made to stand in an upright position while their feet were in apposition. The values for the measurements were recorded to the nearest 0.1cm and the mean values calculated from three measurements.

### Assessment of blood pressure

The blood pressure (BP) of the participants was also measured. Each participant was made to take 10 minutes of rest before the BP measurement was done. In order to ensure consistency and to obtain reliable and true BP based on recommendation by Pickering et al., 2005, the measurement was done from the left arm of all the participants using a digital sphygmomanometer (OMRON, Japan). Appropriate size cuff was placed on the left upper arm. Both Systolic Blood Pressure (SBP) and Diastolic blood pressure (DBP) were taken in triplicates at five minutes intervals. Blood pressure was categorized as: Normal-SBP < 120mmHg and DBP < 90mmHg. Hypertension-SBP > 140mmHg and DBP > 90mmHg (WHO, 1999).

### Dietary assessment

Information on food consumption was obtained using a validated food frequency questionnaire (FFQ). A 3-Day repeated 24hour dietary recall, which included a weekend, was also done to assess dietary intake. Participants were aided to describe all meals and fluid consumed the previous day and to estimate quantities consumed using commonly used household handy measures in Ghana. The quantities of intake were converted to gram equivalents and amounts of nutrients determined from grams of food intake using a template based on Ghanaian foods. Protein, fat, and carbohydrates were calculated as a percentage of total energy intakes.

### Biochemical assessments

Blood Biochemical markers measured from blood serum were serum Total Cholesterol (TC), Triglycerides (TG), High-Density Lipoprotein (HDL), Low-Density Lipoprotein (LDL) and Fasting Blood Glucose, as well as glycated haemoglobin (HbA1c). Before the participants’ venous blood samples were taken from the median cubital vein, they were made to fast overnight for twelve hours. The blood samples were taken and handled by phlebotomists who had been given additional training for the purpose of this study. An Automated Biochemistry Analyzer (Hitachi, 7600-020ISE) was used to run the biochemical test at the Medical Laboratory of Tamale Technical University Hospital in Tamale. The tests were carried out by a Principal Biomedical Scientist.

### Procedure for Biochemical Analysis

**A**bout 7ml of blood sample was collected from via venipuncture from each participant by trained phlebotomist for lipid profile, HbA1c and fasting plasma glucose analysis. Ice chest-containing ice packs were used to temporarily store and transport blood samples from the field to the clinical laboratory of Tamale Technical University for analysis. About 4mls of the blood sample was dispensed into a gel activator tube and the remaining 3mls was dispensed 1.5 ml each into a fluoride tube for fasting plasma glucose (FPG) and an EDTA tube for HbA1c tests. Serum was obtained from the blood samples in the gel activated tube by centrifuging for ten minutes using an eppendorf centrifuge 5804 at 4000 rotations per minute (rpm). Test kits from Medsource Ozone Biomedicals Pvt were used for all the biochemical tests.

### Principle of Fasting Plasma Glucose

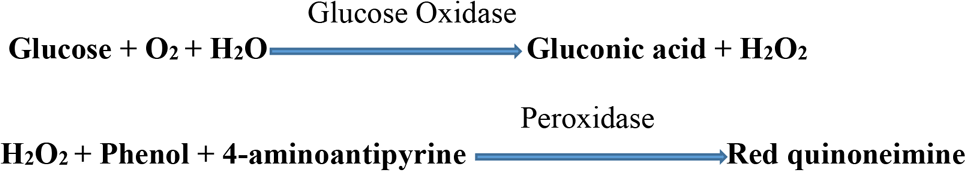

Glucose oxidase oxidizes glucose present in the plasma to yield gluconic acid and hydrogen peroxide which subsequently reacts with hydroxybenzoate and 4-aminoantipyrine in the presence of peroxide enzyme to give a red-violet quinoneimime dye.

### Method

Blood samples in the fluoride tubes were allowed to stand for about 10 minutes. Then the sampled were centrifuged for about 5 minutes. About 10µL of plasma obtained after centrifugation was pipetted into labelled test tubes and 1ml of glucose reagent added. The test tubes were incubated in a water bath at 37oC for about 1o minutes. A blank was then prepared by pipetting about 10µL of deionized water into a labelled test tube and 1ml of glucose reagent added to it. The absorbance of the solution was read at 510nm using a semi-automated spectrophotometer (Biolabo Diagnostic Kenza Biochemistry Try, France) to determine blood glucose concentration.

### Glycated Haemoglobin (HbA1c) Test Principle

Haemolyzed whole blood is mixed with weak binding cation-exchanging resin continuously for five (5) minutes. There is adherence to non-glycated haemoglobin to resin during mixing and the glycated haemoglobin is detached in the precipitate. The precipitate binding the glycated haemoglobin is divided from the resin by mixing and filtering. The ratio of absorbance of glycated haemoglobin fraction to total haemoglobin fraction is measured to determine the percentage of glycated haemoglobin (Gabbay et al., 1977; Gonen and Rubenstein, 1978).

## Methods

### Preparation of haemolyte

About 250 µL of lysing reagent was pipetted into labelled sample tubes, then, about 50 µL of properly mixed whole blood was added and incubated at room temperature for complete lysis of erythrocytes to occur.

### Preparation of glycated haemoglobin

The toppers of prefilled resin tubes were removed and about 100 µL of haemolysate was added to the resin tubes. The resin separators were inserted in a way that allows the rubber sleeve to be about 1cm above resin suspension surface. The haemolysate in the tubes was mixed for five continuous minutes on a stirrer and afterwards, the resin separators were inserted to firmly pack resin at the bottom for the supernatant to enter the separator tubes. The supernatant was transferred into a cuvette and absorbance was at 415 nm using distilled water as blank.

### Preparation of total haemoglobin

An automated pipette was used to measure deionized water into previously labelled test tubes. Then about 20 µL of haemolysate was added to each test tube and properly mixed. Absorbance was read at 415 nm against distilled water blank. Percentage of glycated haemoglobin (GHA1%) was determined using the formular below:

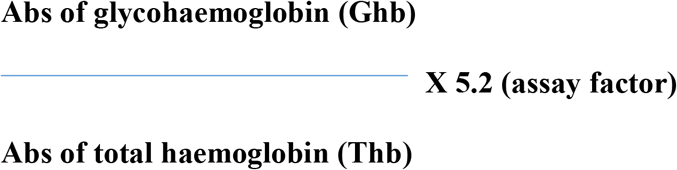

Using a conversion table, percentage glycated haemogloin A1c (HbA1c) that corresponds with the % GHbA1 obtained from the calculation above was obtained.

### Lipid Profile (Serum Triglyceride-TG, Total Cholesterol-TC, Low Density Lipoprotein-Cholesterol-LDL-C, High Density Lipoprotein Cholesterol-HDL-C)

**G**el and clot activator tubes with about 4ml of blood samples were allowed to stand for about 30 minutes after centrifuging for about 10 minutes at 4000rpm to obtain serum. The centrifugation was done using the Eppendorf Centrifuge 5804, EpperdortNetherler-Hinz, Germany. Lipid profile parameters were then determined from the serum using reagents from Forttress Diagnostics Limited, UK.

### Serum Triglycerides Test Principle

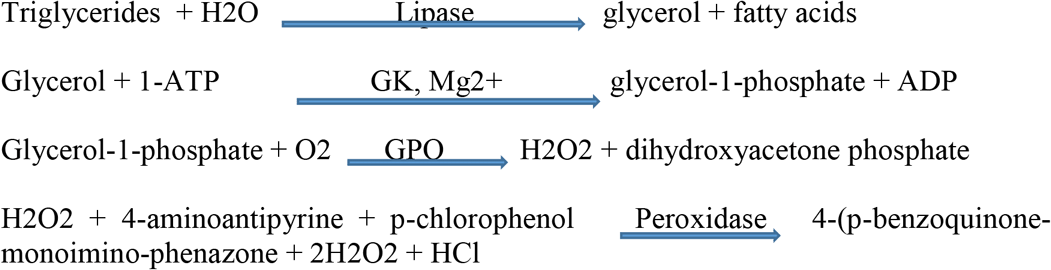

**Note: GPO-glycerol phosphate oxide, GK-glycerol kinase**

### Method for Determining Serum Triglycerides (TG) Concentration

A sterile automatic micropipette was used to pipette 10µL of serum into a labelled sterile test tube and 1000 µL of triglyceride reagent added. The test tube was gently stirred and incubated for 5 minutes in a 37°C water bath. The concentrations of the triglycerides were determined by reading the absorbent of the red colour dye at 505nm with 1 cm light path cuvette in a semi-automated spectrophotometer.

### Total Cholesterol Test Principle

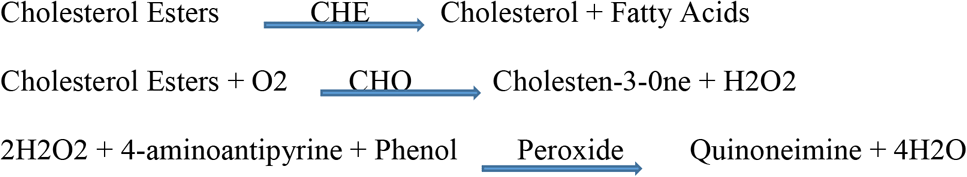

**Note: CHE- Cholesterol Estate CHO- Cholesterol Oxidate**

### Method for Determining Total Cholesterol Concentration

A sterile micropipette was used to pipette about 10µL of serum into a labelled sterile test tube and 1000 µL of cholesterol reagent was added. Another 10 µL of deionized water was put in another test tube and 1000 µL of cholesterol also added to constitute the blank solution. The content of the test tubes were stirred gently and the test tubes were incubated in a 37oC water bath for 5 minutes and then an absorbance was read at 500nm using a semi-automated spectrophotometer.

### Determination of Serum HDL-C Concentration

The determination of serum HDL-C was done using a semi-micro assay. With a sterile automatic micropipette, about 200 µL of serum was transferred into a labelled sterile test tube and about 500 µL of HDL precipitant reagent was added. LDL-C became precipitated from the serum leaving thw HDL-C as supernatant after adding phosphotungstic acid and magnesium chloride containing reagents. The test tubes were incubated at room temperature for 10 minutes after gentle mixing, then centrifugation was was done at 4000 rpm for 10 minutes in an Eppendorf Centrifuge. The CHOD-PAP method was then used to estimate HDL-C content. A blank solution was then prepared by pipetting about 100 µL of supernatant serum HDL into sterile test tubes, then 1ml of HDL reagent was added. The test tubes were gently mixed and incubated at 37oC in a water bath for 5 minutes. HDL-C cpncentration was then determined by measuring absorbance at 500 nm using a semi-automated spectrophotometer.

### Serum LDL-C Concentration

The LDL cholesterol concentration of all the samples was computed using the Friedewald formular: **LDL-C = TC – [HDL-C + (TG/2.2)] mmol/l**

### Assessment of Serum Vitamin D and Determination of Vitamin D Status

Serum 25-hydroxyvitamin D concentration was determined by electrochemiluminescence immunoassay (Leino et al., 2008) using a Roche Modular E170 Analyzer (Roche Diagnostics GmbH) at the Tamale Teaching Hospital. Participants whose serum 25-hydroxyvitamin D levels were 50nmol/L (20ng/ml)-125nmol/L (50ng/ml); 25 nmol/L (10ng/mL) to 50nmol/L (20ng/ml); and < 25nmol/L (<10ng/ml) were considered as having sufficient; insufficient and deficient vitamin D levels respectively (Arundel, et al., 2012).

### Definitions of Anthropometric and Biochemical Parameters

1. **Generalized Obesity (BMI) and Central Obesity (WC)** ➢**WC** ≥ 102 cm for men and ≥ 88 cm for women ➢**Overweight: BMI** 25.0 – 29.9 kg/m2 **Obesity: BMI** ≥ 30 kg/m2 ➢**Underweight: BMI < 18.5kg/m2; Normal weight: 18.5 – 24.9kg/m2** (WHO, 2008: WHO, 2006) ➢**WHR > 0.9 for males and > 0.85 (**WHO, 2016)
2. **Type 2 Diabetes:** ➢FPG ≥ 7mmol/l or HbA1c ≥ 6.5% (WHO 2006)
3. **Serum Lipid Profile** ➢Elevated triglycerides ≥ 1.7mmol/l; ➢Elevated total cholesterol ≥ 6.5mmol/l ➢Elevated LDL ≥ 4.1 ➢Decreased HDL < 1.03mmol/l for men and < 1.3 for females **(NCEP ATP III, 2001)**
4. **Blood Pressure** ➢Normal Blood Pressure: SBP 120 – 140 mmHg ➢High Blood Pressure: SBP > 140 mmHg and DBP > 90mmHg (**WHO, 1999**)
5. **Coronary Risk** ➢High Coronary Risk: TC/HDL-C ≥ 3.5 ➢Low Coronary Risk: TC/HDL-C < 3.5

### Diagnosis of Cardiometabolic Disease (CMD)

The diagnosis of CMD in this study was in accordance with the criteria provided by the WHO (WHO 2007). **Central obesity** was **defined** according to the WHO criteria: WC ≥ 102 cm for men and ≥ 88 cm for women or Waist-to-Hip Ratio (WHR) ≥ 0.90 in men and ≥ 0.85 in women and a WHTR of > 0.50; Plus, any two of the following; raised triglycerides (>1.7mmol/L); reduced HDL-cholesterol (<1.03mmol/L in men and <1.29 mmol/L in women); raised blood pressure (systolic >140 mmHg or diastolic >90 mmHg); raised fasting plasma glucose (≥ 5.6 mmol/L).

### Ethical Consideration

This study was approved by the Council for Scientific and Industrial Research (CSIR) Institutional Review Board (IRB) (Ref: RPN 003/CSIR-IRB/2018). In addition, this study was approved by the Metropolitan Director of Health Services, Tamale (TMHD/MPHs/09). All participants signed informed consent prior to their participation.

### Statistical Analysis

Data was entered into Microsoft excel and analyzed using Statistical Package for Social Sciences (SPSS), IBM Corporation, USA, version 25. The variables were tested for normality by the Kolmogorov-Smirnov Test. Socio-demographic characteristics, anthropometric and biochemical parameters were analyzed using descriptive statistics and expressed as percentages, frequencies, means and standard deviations. Means of continuous variables were compared using one-way analysis of variants (ANOVA). Correlation analysis was performed to determine associations between metabolic factors and vitamin D. The data were expressed as mean ± SD or median (inter-quartile range). T-test was performed to compare two groups of data with normal distribution, while Mann-Whitney U test was performed to compare two or more groups of the data with non-normal distribution. Binary regression analysis was used for evaluating the effects of the variables of CMD on the serum vitamin D status. Multivariate logistic regression analysis was used for evaluating the effects of vitamin D and other risk factors of Cardiometabolic Disease (CMD). A *p*-value of less than 0.050 was considered as statistically significant.

## Results

### Prevalence of CMD risk factors

The mean age of the 302 participants was 38.78 (±10.42) years, similar between the two genders. Table 1 presents summary of risk factors for CMD. The females recorded significantly (p=0.006) higher mean BMI (24.31**±** 7.96) than the males (22.52**±** 3.07), as well as total % body fat (males=17.60%**±**7.90%, females=32.09%**±**9.50%, p<0.001), waist (males=80.74**±** 9.80, females=85.00**±**12.27, p=0.002) and hip circumferences (males=93.45, **±**9.34, females=96.89**±**12.10, p=0.011). On the other hand the males recorded higher serum lipid parameters, such as mean serum total cholesterol (5.74**±** 1.02) and LDL (3.57 **±**1.02) than the females (TC=5.40**±**1.05, p=0.008; LDL=3.22**±**1.09, p=0.005). The mean ages, SBP, DBP, FBG, HbA1c, triglycerides, HDL and visceral fat were similar between males and females.

**Table 1:** Blood glucose, blood pressure, lipid profile and body composition of participants

| Variables | Combined |  | Male |  | Female |  | *p*-value |
| --- | --- | --- | --- | --- | --- | --- | --- |
|  | N | Mean± SD | N | Mean± SD | N | Mean± SD |  |
| Age | 302 | 38.28±**10.9** | 116 | 37.53 ± **11.6** | 186 | 38.75 ± **10.4** | 0.343 |
| SBP(mmHg) | 302 | 134.63±47.97 | 116 | 130.22±37.97 | 186 | 139.04± 57.97 | .108 |
| DBP(mmHg) | 302 | 90.31±33.89 | 116 | 88.85 ±31.92 | 186 | 91.77± 35.86 | .474 |
| Glucose (mmol/l) | 302 | 4.9±1.62 | 115 | 4.83± 1.44 | 185 | 4.92± 1.81 | .652 |
| HBA1C (%) | 302 | 5.27±.57 | 114 | 5.28± .56 | 182 | 5.25± .57 | .686 |
| Serum TG (mg/dl) | 302 | 1.04±.47 | 115 | 1.01±.48 | 185 | 1.06±0.46 | .370 |
| Serum cholesterol | 302 | 5.57±1.04 | 114 | 5.74± 1.02 | 185 | 5.40±1.05 | **.008** |
| LDL (mg/dl) | 302 | 3.34±1.06 | 115 | 3.57, ±1.02 | 185 | 3.22±1.09 | **.005** |
| HDL (mg/dl) | 302 | 1.73±0.22 | 115 | 1.72, ± .23 | 185 | 1.74 ± .20 | .476 |
| BMI (kg/m2) | 302 | 23.42±5.5 | 116 | 22.52± 3.07 | 184 | 24.31± 7.96 | **.006** |
| Body fat (%) | 302 | 24.8%±8.7% | 115 | 17.60%±7.90% | 186 | 32.09%±9.50% | **<0.001** |
| Visceral fat (%) | 302 | 5.53±3.14 | 116 | 5.40±3.17 | 185 | 5.66, ±3.11 | 0.479 |
| WC (cm) | 302 | 82.9±11.0 | 112 | 80.74 ± 9.80 | 183 | 85.00, ±12.27 | **0.002** |
| HC (cm) | 302 | 95.17±10.72 | 112 | 93.45±9.34 | 183 | 96.89, ±12.10 | **0.011** |
| WHR (cm) | 302 | 0.89±0.33 | 99 | 0.86± .08 | 167 | 0.92, ±.57 | 0.291 |

\***Bold figures show that there are significant differences in those variables between males and females.**SD: standard deviation; WC: waist circumference; HC: hip circumference; SBP: systolic blood pressure; DBP: diastolic blood pressure; Glu: fasting glucose; TG: triglyceride; BMI: body mass index; WHR: waist circumference/hip circumference; TC: total cholesterol; LDL: low density lipoprotein cholesterol; HDL: high density lipoprotein.

### Cardiometabolic Disease Prevalence

Figure 1 shows the prevalence of CMD, determined by being centrally obese, plus two CMD risk factors. Of the total population, 6.4% had CMD, all women. Of the female population alone, 10.2% had CMD.

**Fig 1:**
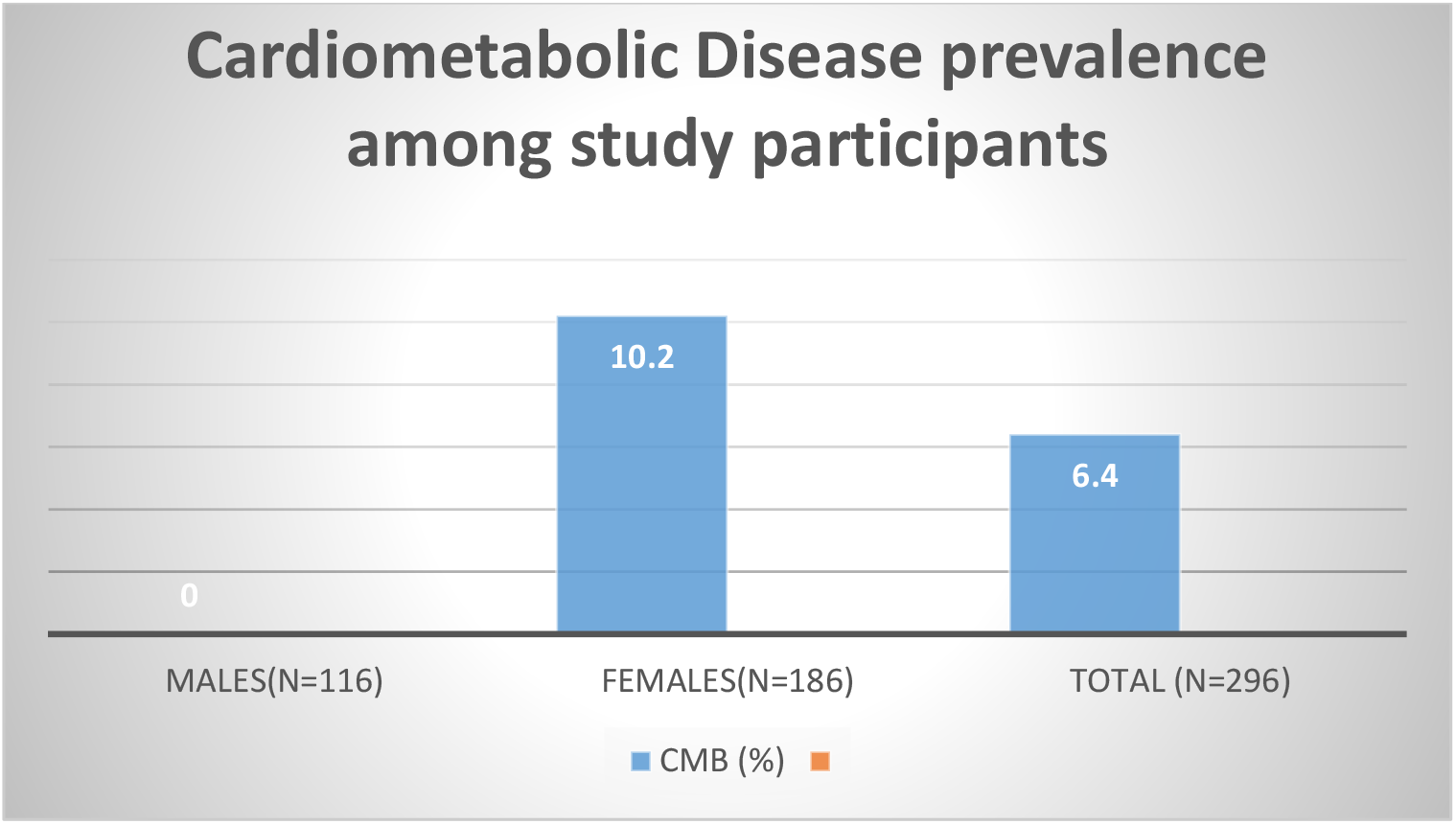
Prevalence of CMD among Study Participants.

### Vitamin D levels and status by age and gender of participants

The distribution of serum vitamin D status is shown in Table 2 below. Seven percent (7%) of the participants had vitamin D deficiency (**<25 nmol/L), 7.5% in the females and 6.5% among the males (p=0.349)**. The mean (±SD) vitamin D levels of those with vitamin D deficiency was 20.1 **±** 2.4 **nmol/L**; that of vitamin D insufficiency was 41.2 ± 7.2 **nmol/L** and vitamin D sufficiency was 80.2 ±17.1 **nmol/L**. The mean (±SD) vitamin D for the study group was 65.2 nmol/L, 66.6 (± 25.3 for males and 63.8 (± 25.8) for females, with no statistical significance between the two categories (*p=0*.*349*). Mean serum vitamin D levels for the different vitamin D categories by age group were different (*p=0*.*047*). Vitamin D deficiency increased with age, from 5% in the 20-29 years category to 12% among the 50-59 year olds, while vitamin D sufficiency decreased with age, from 74.4% in the 20-29 year olds to 48% among the 50-59 years category.

**Table 2:** Serum **25(OH)D**/nmol/L levels and status by gender and age of participants

| S-Vit D/nmol/L | Total | <25<br>nmol/L(Deficiency) | 25-50nmol/L<br>(Insufficiency) | >50nmol/L(Sufficiency) | P value |
| --- | --- | --- | --- | --- | --- |
| Means by gender |  |  |  |  |  |
| Males(mean±SD) | 66.6 ± 25.3 | 19.9±2.7 | 42.5 ± 5.6 | 79.9±18.2 | p=0.349 |
| Females (mean±SD) | 63.8± 25.8 | 20.2 ±2.3 | 40.4 ± 7.8 | 80.3±16.4 |  |
| Total (mean±SD) | 6 | 20.1 ±2.4 | 41.2 ± 7.2 | 80.2±17.1 |  |
| Status by gender |  |  |  |  |  |
| Males N(%) | 116 (38.4%) | 7 (6%) | 30 (25.9%) | 79 (68.1%) | 0.349 |
| Females N(%) | 186 (61.6%) | 14 (7.5%) | 56 (30.1%) | 116 (62.4%) |  |
| Total N(%) | 302 (100) | 21 (7.0%) | 86 (28.5%) | 195 (64.6%) |  |
| Means by age |  |  |  |  |  |
| 20-29 mean (±SD) | 69.9±25.3 | 20.7 ± 2.5 | 41.1 ± 7.8 | 81.2 ± 17.8 | 0.047 |
| 30-39 mean (±SD) | 66.2±23.9 | 18.7 ± 2.8 | 41.2 ± 7.6 | 78.9 ± 15.5 |  |
| 40-49 mean (±SD) | 59.6±23.8 | 20.7 ± 2.6 | 42.0± 6.8 | 75.4 ± 16.7 |  |
| 50-59 mean (±SD) | 59.3±28.8 | 20.2 ± 1.8 | 39.6 ± 7.5 | 85.4 ± 16.6 |  |
| 60-69 mean (±SD) | 72.3±28.6 | 0 | 43.8 ± 3.1 | 88.2 ± 23.1 |  |
| Status by age |  |  |  |  |  |
| 20-29 N(%) | 78 | 4 (5.1%) | 16 (20.5%) | 58 (74.4%) | 0.047 |
| 30-39 N(%) | 95 | 5 (5.3%) | 24 (25.3%) | 66 (69.5%) |  |
| 40-49 N(%) | 65 | 6 (9.2%) | 21 (32.3%) | 35 (58.5%) |  |
| 50-59 N(%) | 50 | 6 (12%) | 20 (40%) | 24 (48%) |  |
| 60-69 N(%) | 14 | 0 | 5 (35.7%) | 9 (64.3%) |  |

### Relationship between Serum Vitamin D and CMD Risk Factors

Table 3 compares the mean CMD risk factors between participants with vitamin D deficiency, insufficiency and sufficiency. Apart from serum total cholesterol, HDL and triglycerides, all the CMD risk factors were different between the different vitamin D status categories. Mean age of those with VDD is significantly higher than those with VDS (p=0.009). Both mean SBP (P<0.001) and DBP (P<0.001) increased with VDD, with the least in the VDS, followed by VDI and the highest in the VDD. Consistently, the means for the following: FBG (p=0.032), BMI (p=0.004), HbA1c (p=0.040), and LDL (p=0.047) are highest in the VDD group, followed by the VDI group and lowest in the VDS group. Therefore, all the CMD risk factors seem to increase with vitamin D deficiency. Similarly, all the body composition parameters, being mean visceral fat (p=0.003), percent total body fat p<0.001), WC (p=0.007) and WHR (p=0.002) are all significantly highest in the VDD group and lowest in the VDS group, with the exception of HC. CMD also increased with vitamin D deficiency. Close to 30% of participants with vitamin D deficiency had CMD compared to 5.85 in the vitamin D insufficiency and 4.1% in the vitamin sufficiency group (p<0.001). Overweight and obesity (using BMI cutoffs) were also higher among the vitamin D deficiency group (47.6%) and lowest in the vitamin sufficiency group (18.46%) (p<0.001). Similarly, more than 4 in 10 participants with central obesity were vitamin D deficient, compared with about 3 in 10 in the vitamin D insufficiency group and less than 2 in 10 in the vitamin D sufficient group.

**Table 3:** Comparison of variables in different Serum-Vitamin D/nmol/L levels

| Variables | N | Vitamin D Deficiency |  | N | Vitamin D Insufficiency |  | N | Vitamin D Sufficiency |  | *p*-value |
| --- | --- | --- | --- | --- | --- | --- | --- | --- | --- | --- |
|  |  | Mean | SD |  | Mean | SD |  | Mean | SD |  |
| Age (y) | 21 | 40.43 | 9.77 | 86 | 40.95 | 11.17 | 195 | 36.875 | 10.64 | **0.009** |
| SBP (mmHg) | 21 | 177.13 | 69.54 | 86 | 158.43 | 70.95 | 195 | 121.14 | 25.72 | **<0.001** |
| DBP (mmHg) | 21 | 114.42 | 50.49 | 86 | 102.10 | 41.95 | 195 | 83.04 | 24.94 | **<0.001** |
| FBG | 21 | 5.58 | 2.01 | 86 | 5.09 | 2.17 | 193 | 4.71 | 1.33 | **0.032** |
| HBA1C | 21 | 5.21 | 0.55 | 85 | 5.40 | 0.62 | 190 | 5.21 | 0.54 | **0.040** |
| LDL | 21 | 3.91 | 1.24 | 86 | 3.34 | 1.08 | 193 | 3.30 | 1.05 | **0.047** |
| BMI (kg/m2) | 21 | 27.58 | 11.13 | 86 | 24.25 | 4.73 | 193 | 22.90 | 6.48 | **0.004** |
| Body fat (%) | 21 | 32.56 | 11.82 | 86 | 29.73 | 12.06 | 194 | 24.50 | 10.46 | **0.001** |
| Visceral fat | 21 | 7.05 | 2.97 | 86 | 6.11 | 3.04 | 194 | 5.16 | 3.11 | **0.005** |
| WC (cm) | 21 | 88.31 | 12.32 | 83 | 85.57 | 12.63 | 191 | 81.89 | 10.72 | **0.007** |
| WHR (cm) | 19 | 1.25 | 1.69 | 71 | 0.87 | 0.11 | 176 | 0.86 | 0.077 | **0.002** |
| Overweight + obesity (%) | 10 | 47.62 |  | 36 | 41.86 |  | 36 | 18.46 |  | **<0.001** |
| Abdominal obesity (%) | 9 | 42.86 % |  | 27 | 31.40 |  | 37 | 18.97 |  | **<0.001** |
| CMD | 6 | 28.6 |  | 5 | 5.8 |  | 8 | 4.1 |  | **<0.001** |
| Serum triglycerides | 21 | 1.043 | 0.46 | 86 | 1.01 | 0.42 | 193 | 1.05 | 0.49 | 0.837 |
| Serum cholesterol | 21 | 5.97 | 1.22 | 86 | 5.53 | 1.05 | 192 | 5.47 | 1.02 | 0.120 |
| HDL | 21 | 1.68 | 0.18 | 86 | 1.73 | 0.20 | 193 | 1.74 | 0.22 | 0.517 |
| HC (cm) | 21 | 96.65 | 21.96 | 83 | 97.30 | 11.10 | 191 | 94.73 | 9.45 | 0.200 |

WC: waist circumference; HC: hip circumference; SBP: systolic blood pressure; DBP: diastolic blood pressure; Glu: fasting glucose; TG: triglyceride; BMI: body mass index; WHR: waist circumference/hip circumference; TC: total cholesterol; LDL: low density lipoprotein cholesterol; HDL: high density lipoprotein cholesterol.

### Binary Regression Analysis to Predict Vitamin D and C Risk

A binary regression (logistic regression) was performed to ascertain the effects of Age, Gender, SBP, DBP, FPG, HbA1c, STG, SC, LDL, HDL, BF, WC, HC, WHR, Visceral fat, and BMI on the likelihood that a participant or respondent’s CMD risk factor will have an influence on vitamin D deficiency. The logistic regression model was statistically significant, *χ*^2^ =40.589, *p* <0.001. The model explained 36.4% (Nagelkerke R Square) of the variance in Vitamin D deficiency levels and correctly classified 91.4% of cases. From the results in Table 4, SBP (p = 0.003) and FBS (p = 0.002) added significantly to the model/prediction. The information in the “Variables in the Equation” table can be used to predict the probability of an event occurring based on a one-unit change in an independent variable when all other independent variables are kept constant. As such the table shows that a unit increase in SBP and FPG were associated with a decrease likelihood of exhibiting Vitamin D deficiency levels. The odds of having Vitamin D deficiency are 0.055 (5.5%) times greater in participants with high SBP whilst the odds of having Vitamin D deficiency are 0.076 (7.6%) times greater in participants with high FPG. The results from the binary regression analysis showed that participants with normal SBP had reduced odds (OR 0.055, 95 CI) for vitamin D deficiency compared with those with high SBP. Also, those with low FBS had reduced odds (0.076, 95 CI) for vitamin D deficiency compared with those with high FBS or diabetics. For LDL, those with normal LDL levels had reduced odds for vitamin D (OR 0.362 95 CI) compared with those with high LDL levels.

**Table 4:** Binary Regression Analysis to Predict Vitamin D and CMD Risk

| Covariates | Categories | Odds Ratio (OR) | P-value | Confidence Interval |  |
| --- | --- | --- | --- | --- | --- |
|  |  |  |  | Lower limit | Higher limit |
| Age |  | 1.024 | .397 | .970 | 1.080 |
| Gender | Female | 1.524 | .567 | .361 | 6.435 |
|  | Male | 1 |  |  |  |
| Systolic Blood Pressure | High | .055 | **.003** | .008 | .361 |
|  | Normal | 1 |  |  |  |
| Diastolic Blood Pressure | High | 1.192 | .837 | .224 | 6.346 |
|  | Normal | 1 |  |  |  |
| Fasting Blood Glucose | High | .076 | **.002** | .014 | .404 |
|  | Normal | 1 |  |  |  |
| HbA1c | High | 8.292 | .146 | .478 | 143.879 |
|  | Normal | 1 |  |  |  |
| Serum Triglyceride | High | .661 | .729 | .064 | 6.868 |
|  | Normal | 1 |  |  |  |
| Serum Cholesterol | High | .1.292 | .799 | .179 | 9.319 |
|  | Normal | 1 |  |  |  |
| Serum LDL | High | .362 | .088 | .112 | 1.163 |
|  | Normal | 1 |  |  |  |
| Serum HDL | High | 1 |  |  |  |
|  | Low | 1.175 | .797 | .342 | 4.036 |
| BF(1) | High | 4.547 | .251 | .343 | 60.276 |
|  | Low | 1 |  |  |  |
| WC1(1) | High | .435 | .180 | .129 | 1.468 |
|  | Normal | 1 |  |  |  |
| HC2(1) | High | 1.292 | .684 | .377 | 4.428 |
|  | Low | 1 |  |  |  |
| WHR1(1) | High | .736 | .651 | .196 | 2.768 |
|  | Normal | 1 |  |  |  |
| Visceralfat1 (1) | High | .530 | .623 | .042 | 6.660 |
|  | Normal | 1 |  |  |  |
| BMI6(1) | Obese | 1.518 | .542 | .397 | 5.811. |
|  | Normal | 1 |  |  |  |
| Constant |  | 7.605E+16 | .999 |  |  |

WC: waist circumference; HC: hip circumference; SBP: systolic blood pressure; DBP: diastolic blood pressure; Glu: fasting glucose; TG: triglyceride; BMI: body mass index; WHR: waist circumference/hip circumference; TC: total cholesterol; LDL: low density lipoprotein cholesterol; HDL: high density lipoprotein cholesterol.

**Table 5:** Multiple logistic regression analysis for CMD

|  | B | S.E. | *p*-value | OR | 95% C.I.for OR |  |
| --- | --- | --- | --- | --- | --- | --- |
|  |  |  |  |  | Lower | Upper |
| FBS | .301 | .129 | 0.020 | 1.351 | 1.049 | 1.739 |
| Serum TG | 1.722 | .696 | 0.013 | 5.596 | 1.429 | 21.909 |
| LDL | 1.887 | .454 | <0.001 | 6.597 | 2.709 | 16.063 |
| WC | .194 | .040 | <0.001 | 1.214 | 1.123 | 1.312 |
| Constant | -31.628 | 5.769 | <0.001 | .000 |  |  |

### Predictors of Cardiometabolic Disease (CMD)

From multivariate analysis, filtering FBS, Serum TG, LDL and WC filtered out by in-and-out stepwise method showed FPG, Serum Triglycerides, LDL, and WC as independent risk factors of CMD with p-values of 0.020, 0.013, 0.001 and 0.001 respectively.

The dependent variable (CMD), was categorized into two; those who have CMD were assigned 1, and those who do not assigned 0. As such the dependent variable (CMD) is dichotomous (binary). Logistic regression is the appropriate regression analysis to conduct when the dependent variable is dichotomous (binary). Logistic regression is used to describe data to explain the relationship between one dependent binary variable and one or more nominal, ordinal interval or ratic level independent variables. With regards to this study, we are interested in how the independent variables: BP, WC, Body fat, visceral fat, Serum TG, FPG, LDL, and obesity have influenced the probability of having CMD. In applying the logistic regression, the main assumption of no multicollinearity, no outliers and dependent variables been dichotomous was checked.

## Discussion

This study was conducted to look at the interrelationship between serum vitamin D status and risk factors of cardiometabolic disease (CMD) such as obesity, diabetes, hypertension, and dyslipidemia among apparently healthy adults of northern extraction (Dagombas) in Tamale.

In this study, just 7% of the participants were vitamin D deficient, while 28.5% were vitamin D insufficient. Thus, a total of 35.5% of the participants were either Vitamin D deficient or insufficient. We found the mean levels of serum vitamin D among those deficient to be about twice less in the vitamin D insufficient and four-fold less in the vitamin D sufficient group, irrespective of their gender. Our study reported a very low proportion of vitamin D deficiency, compared to what other studies have reported among Ghanaian participants. A study conducted among 92 pre-and post-menopausal women with T2DM aged more than or equal 25 years reported prevalence of hypovitaminosis D of 92.2% (Linda A. F. et. al., 2018). Another study conducted in diabetics reported 92.4% among T2DM participants and 60.2% in non-diabetics controls (Fondio et. al., 2017). Unlike these studies, in which the subjects were diabetics, this current study was conducted among healthy participants, who did not show any signs and symptoms of any chronic disease. A similar study conducted among healthy blood donors reported a vitamin D prevalence rate of 43.6%. Comparing the prevalence of hypovitaminosis D among the two other studies in which the participants are diabetics, the prevalence in both studies reported over 90% prevalence compared with 43.6% prevalence among the healthy blood donors and about 7% and 35.5% of deficiency and insufficiency in this current study. Therefore, the high prevalence of hypovitaminosis among diabetics strongly implicates hypovitaminosis D as a risk factor of T2DM and its consequent effects on the other CMD risk factors.

In the current study, vitamin D status in males and females are of the same levels, which is contrary to the findings by Bolland et al., 2006; Lu et al., 2012; and Borissonva et al., 2013 in which the vitamin D status was found to be lower in males than in females. However, the current study found more of the female participants being overweight and obese than their male counterparts. Therefore, the adipose tissue effect on vitamin D status, in which case women tend to have more adipose tissue with consequential effect on their vitamin D levels, compared to males is not apparent in the findings of this study. For instance, Frisch, 1994; Wortsman et al., 2000, and supported by Rosenstreich et al., 1971 reported that women with high adipose tissue tend to have hypovitaminosis D and that adipose tissue effect on vitamin D concentration accounted for low vitamin D levels in women compared with their male counterparts. The results also showed vitamin D deficiency increased with age and significant differences in serum 25 (OH)D levels among males and females in the different age groups (p=0.047). The female participants in this age group had lower concentration of serum 25 (OH)D levels (52.7nmol/l) compared with the male counterpart who had 93.05nmol/l of serum vitamin D concentration. This finding is in agreement with established knowledge as reported by Maclaughlin & Holick, 1985 that synthesis of vitamin D is markedly decreased in the elderly compared to the younger population.

Cardiometabolic disease was less than 10% in the combined population but all those with CMD were women, representing more than 10%. Being a healthy population, we found this observation worrying, but even more so as this was only reported among the females. Our findings showed that the risk factors for CMD were FBG, serum TG, LDL and waist circumference. This implies that for this population, the key risk factors that should be watched carefully are central obesity, dyslipidaemia and hyperglycaemia. In terms of the CMD risk factors, the men had more dyslipidaemia while the women had more central obesity and BMI derangements. Thus, although none of the men had CMD, the presence of dyslipidaemia among the men suggests they are equally on their way to developing CMD.

Apart from serum total cholesterol, HDL and triglycerides, all the CMD risk factors were different between the different vitamin D status categories. Mean age of those with VDD is significantly higher than those with VDS. Both mean SBP and DBP increased with VDD. Consistently the means for FBG, BMI, HbA1c and LDL were highest in the VDD group and lowest in the VDS group. Therefore, all the CMD risk factors increased with vitamin D deficiency. Similar observations are reported for all the body composition visceral fat, percent total body fat, WC and WHR, whereby all the means were significantly highest in the VDD group and lowest in the VDS group, with the exception of HC. Overweight and obesity, measured by BMI were also highest, at about 50%, among vitamin D participants and less than 2 in 10 in their vitamin sufficiency counterparts. Similarly, more than 4 in 10 participants with central obesity were vitamin D deficient, compared with about 3 in 10 in the vitamin D insufficiency group and less than 2 in 10 in the vitamin D sufficient group. CMD itself was highest in the vitamin D deficient participants (about 30%) compared to 5.85 in the vitamin D insufficiency and 4.1% in the vitamin D sufficiency groups (p<0.001). Thus vitamin D deficiency most likely increased CMD risk. This increased CMD risk is demonstrated by the results of the binary regression analysis of all the risk factors of CMD, which showed that vitamin D status was independently predicted by Systolic BP (p-value of 0.002), fasting blood sugar (FBS) levels (0.001), and LDL-cholesterol (p-value of 0.048).

Apart from the contrasting findings between gender and vitamin D status (Palacios *et al*., 2012 and Vierucci *et al*., 2013), most study findings are in line with our findings (Rockell *et al*., 2005; Hill et al., 2008; and Janssen *et al*., 2013). Increased fasting blood glucose with its associated insulin resistance (IR), are known to play significant roles in the pathophysiology of metabolic diseases (Fujita, 2007). For vitamin D and blood pressure, study findings have been mixed. Most of the cross-sectional studies (Hintzpeter et al., 2007; Scragg et al., 2007; Hyppönen *et al*., 2008; Forman et al., 2013; and Nasri et al., 2014) reported that deficiency or insufficiency of vitamin D was associated with increased BP, as we found, while, Snijder *et al*. (2007) and Reis *et al*. (2007) found no inverse relationship. Regarding body weight and fat and vitamin D status, Wortsman et al., (2000), found that obese respondents reported 57% lower levels in serum vitamin D relative to age-matched non-obese counterparts, after the two groups were exposed to equal amounts of ultraviolet radiations. Wortsman et al., 2000; Drincic et al., 2012; and Rock *et al*. (2012) also reported inverse relationships between body size and serum vitamin D status. Arunabh et al., (2003) and Blum et al., (2008), reported BMI and fat content in non-obese people was negatively correlated with vitamin D levels. Our findings between CMD itself and vitamin D status have also been reported (Jorde & Grimnnes, 2011; Renzaho et al., 2011; Dolinsky et al., 2013; Saneei et al., 2013). All the participants in this study with serum vitamin D sufficiency did not have risk factors of cardiometabolic disease, suggesting that vitamin D sufficiency is protective of cardiometabolic disease. Thus, our findings unequivocally confirmed the inverse association between some of the CMD risk factors (SBP, FBS, LDL) and vitamin D status. Also, the results of multiple logistics regression for risk factors of CMD showed statistical significance for FBS, serum TG, LDL and WC with increased odds of CMD for participants with elevated levels of these risk factors compared with those who have normal levels of these risk factors at 95% confidence interval (CI).

## Limitations

The dietary impact on vitamin D status of the study participants was not assessed. This could have significant influence on their serum vitamin D status. Also, this study was not conducted on a large scale due to resource constraints.

## Conclusions

This study has shown that hypovitaminosis D (vitamin D deficiency) contributes to the risk of developing cardiometabolic disease (CMD) by its inverse relationship between SBP, FBS and LDL-cholesterol. SBP, FBS and LDL-cholesterol. These CMD risk factors have all shown statistical significant relationship with vitamin D. Participants with normal SBP, low/normal FBS and normal LDL-cholesterol all had reduced odds for vitamin D deficiency compared with increased odds for the deficiency of vitamin D among those with high levels of these CMD risk factors. Therefore, hypovitaminosis D is implicated in contributing to elevated levels of these risk factors, thereby predisposing to developing CMD. However, the prevalence of hypovitaminosis D was found to be relatively lower than have been reported by other studies. Also, FBS, serum TG, LDL, and WC all independently predicted CMD from a logistics regression analysis of CMD. Therefore, public health interventions aimed at correcting hypovitaminosis D could contribute to reducing the incidence of CMD. Similar studies should be conducted among other ethnic groups in other parts of Ghana to enable a comparison with this present study conducted among participants of dagomba ancestry in the northern region of Ghana. This will enable the impact of ethnic connotations on the association between vitamin D status and the risk factors of CMD to be established for more targeted preventive interventions in curbing the prevalence and incidence of hypovitaminosis D related CMD.

## Data Availability

data will be provided later as per the policy

## Acknowledgements

The authors express thanks to all participants for making this study possible.

## Author Contributions

Conceptualization, A.-M.B, R. A. A K.S.V, B. E and.; methodology, A.-M.B, R. A. A. K.S.V. and S.A.; software, S.A., A. R. H. and A.-M.B.; validation, A.-M.B.; formal analysis, A.-M.B., R. A. A. and A. R. H.; investigation, A.-M.B, and R.A.A.; resources, A.-M.B, K.S.V., R.A.A. and B.E.; data curation, A.-M.B, A. R. H and R.A.A.; writing— original draft preparation, A.-M.B, and R.A.A.; writing—review and editing, A.-M.B, and., R.A.A.,; supervision, R.A.A.; project training and administration, K.S.V., R.A.A., and B.E.; funding acquisition, A.-M.B, R. A. A., K.S.V. and B.E. All authors have read, edited and approved the published version of the manuscript. All authors have read and agreed to the published version of the manuscript.

## Conflicts of Interest

The authors declare no conflict of interest.

## Notes

### Competing Interest Statement

The authors have declared no competing interest.

### Funding Statement

Personal funding as a PhD candidate

### Author Declarations

This study was approved by the Council for Scientific and Industrial Research (CSIR) Institutional Review Board (IRB) (Ref: RPN 003/CSIR-IRB/2018). In addition, this study was approved by the Metropolitan Director of Health Services, Tamale (TMHD/MPHs/09).

